# Polypharmacy and Proton Pump Inhibitor Use Independently Predict One-Year Mortality in Critical COVID-19: An Explainable AI–Based Survival Analysis

**DOI:** 10.1101/2025.10.27.25338863

**Authors:** Malin Hjärtström, Ingrid Didriksson, Martin Spångfors, Hans Friberg, Andreas Jakobsson, Attila Frigyesi

## Abstract

Mortality among patients admitted to intensive care with coronavirus disease 2019 (COVID-19) remains substantial despite advances in management. The contribution of pre-admission medication profiles to long-term survival is poorly defined. We analysed 497 adults with confirmed COVID-19 admitted to six intensive care units in southern Sweden between May 2020 and May 2021. Clinical and laboratory data were combined with prescription information from the national drug registry; drugs dispensed at least twice within eight months before admission were classified by Anatomical Therapeutic Chemical code. Polypharmacy was defined as the use of five or more medications. An XGBoost survival model with a Cox partial-likelihood objective was trained to predict one-year mortality and interpreted using SHapley Additive exPlanations (SHAP). The model achieved a concordance index of 0.74. Age was the strongest predictor of mortality, followed by the number of medications per patient, which ranked above the Charlson Comorbidity Index and Clinical Frailty Scale. Proton pump inhibitors were the only individual drug class among the top predictors, showing a modest positive association with mortality, whereas angiotensin-converting enzyme inhibitors and angiotensin II receptor blockers had negligible contributions. These findings identify cumulative medication burden as an independent and clinically relevant marker of vulnerability in critical COVID-19.

## Introduction

Coronavirus disease 2019 (COVID-19), caused by severe acute respiratory syndrome coronavirus 2 (SARS-CoV-2), was declared a global pandemic in March 2020^1^. Although most infections are mild, a considerable proportion of patients develop severe respiratory failure and multiorgan dysfunction requiring intensive care and invasive mechanical ventilation. During the first pandemic wave, mortality among mechanically ventilated patients exceeded 40%, particularly in older adults and those with pre-existing comorbidities^2^. Although advances in intensive care have improved survival, critically ill COVID-19 patients continue to face a high risk of long-term morbidity and mortality. While vaccination has reduced the number of individuals progressing to critical illness^3^, ICU mortality rates appear comparable between vaccinated and unvaccinated patients^4,5^. This underscores the need to delineate the drivers of mortality in this population.

SARS-CoV-2 infects host cells via membrane-bound angiotensin-converting enzyme 2 (ACE2), which serves as the entry receptor for the viral spike protein. Early in the pandemic, this mechanism prompted speculation that angiotensin-converting enzyme inhibitors (ACEi) or angiotensin II receptor blockers (ARB) might increase susceptibility to infection by upregulating ACE2 expression. An alternative hypothesis suggested potential protection through attenuation of angiotensin II type 1 receptor–mediated vasoconstriction and inflammation during ACE2 depletion^6^. Observational and experimental studies have since largely refuted a harmful association and instead suggested neutral or modestly protective effects on clinical outcomes^7,8^. Nevertheless, uncertainty persists regarding their impact in the most severely affected patients—those requiring intensive care—where haemodynamic instability and acute renal dysfunction frequently complicate treatment decisions.

Beyond renin–angiotensin blockade, polypharmacy and medication burden more generally have emerged as potential modifiers of COVID-19 outcomes. Polypharmacy reflects multimorbidity, frailty, and increased vulnerability to drug–drug and drug–disease interactions, each of which may exacerbate the pathophysiology of critical illness. Prior analyses within the same regional intensive care network in southern Sweden demonstrated that age, frailty, systemic inflammation, and intensive care unit (ICU) strain were principal drivers of mortality in critical COVID-19^2^. Whether long-term medication profiles provide independent prognostic information beyond these factors remains unknown.

Proton pump inhibitors (PPIs) are of particular interest because several cohort and meta-analytic studies have linked chronic PPI therapy to increased risks of pneumonia, enteric infection, and adverse outcomes in COVID-19^9,10^. Conversely, other drug classes such as calcium-channel blockers, *β*-blockers, and dipeptidyl peptidase-4 inhibitors have been hypothesised to confer modest protection through endothelial or immunomodulatory mechanisms^11–13^. However, interpretation of such pharmacoepidemiological associations is hampered by residual confounding, indication bias, and heterogeneous exposure definitions.

To address these uncertainties, we combined prospectively collected plasma biomarkers from the SWECRIT biobank with high-quality registry data, encompassing patients admitted to intensive care for COVID-19 across Skåne county in southern Sweden. Using machine-learning–enhanced survival analysis, our primary objective was to identify pre-admission medications associated with one-year mortality after ICU admission, with particular focus on ACEi and ARB exposure, polypharmacy, and other common drug classes. A secondary objective was to integrate clinical and biochemical variables to improve understanding of the pathophysiological determinants of long-term outcome in critical COVID-19.

## Results

After exclusion of one individual lacking one-year follow-up data, the final study population comprised 497 critically ill patients with confirmed SARS-CoV-2 infection admitted to six southern Swedish ICUs between May 2020 and May 2021. Of these, 35% (172/497) were complete cases, and 19 of 89 candidate variables (21%) had no missing data. Twelve variables were excluded due to more than 20% missingness. The overall demographic and clinical characteristics for the thirteen top-ranked predictors and ongoing medications are summarised in Table 1. The complete characteristics table is provided in Supplementary Table S1.

**Table 1.** Patient characteristics stratified by one-year mortality from ICU admission.

| Characteristic | Missing | One-year mortality |  | p-value<sup>2</sup> |
| --- | --- | --- | --- | --- |
|  |  | Survivors (N = 235)<sup>1</sup> | Non-survivors (N = 262)<sup>1</sup> |  |
| # Days in hospital before ICU | 2.0% | 2.0 (0.0, 5.0) | 3.0 (1.0, 7.0) | < 0.001 |
| # Meds/Patient | 5.4% | 3.0 (1.0, 7.0) | 5.0 (2.0, 10.0) | < 0.001 |
| A10BA | 5.4% | 42 (19%) | 38 (16%) | 0.4 |
| A10BK | 5.4% | 9 (4.0%) | 10 (4.1%) | > 0.9 |
| A10BJ | 5.4% | 11 (4.8%) | 14 (5.8%) | 0.7 |
| A12AX | 5.4% | 8 (3.5%) | 25 (10%) | 0.004 |
| ACEi | 5.4% | 48 (21%) | 43 (18%) | 0.3 |
| Age (year) | 0.0% | 62 (53, 68) | 70 (61, 75) | < 0.001 |
| Albumin (g/L) | 3.6% | 27.8 (25.2, 29.9) | 27.3 (24.8, 30.0) | 0.2 |
| ARB | 5.4% | 31 (14%) | 61 (25%) | 0.002 |
| B01AC | 5.4% | 31 (14%) | 66 (27%) | < 0.001 |
| B01AF | 5.4% | 10 (4.4%) | 19 (7.8%) | 0.12 |
| B03BA | 5.4% | 23 (10%) | 27 (11%) | 0.7 |
| B03BB | 5.4% | 4 (1.8%) | 18 (7.4%) | 0.004 |
| BMI (kg/m<sup>2</sup>) | 2.4% | 31 (27, 35) | 29 (26, 34) | 0.016 |
| C01DA | 5.4% | 7 (3.1%) | 14 (5.8%) | 0.2 |
| C03CA | 5.4% | 14 (6.2%) | 32 (13%) | 0.011 |
| C07AB | 5.4% | 52 (23%) | 77 (32%) | 0.033 |
| C08CA | 5.4% | 54 (24%) | 59 (24%) | >0.9 |
| C10AA | 5.4% | 51 (22%) | 85 (35%) | 0.003 |
| CCI | 0.0% | 2.00 (1.00, 3.00) | 3.00 (2.00, 4.00) | < 0.001 |
| CFS | 0.8% | 3.00 (2.00, 3.00) | 3.00 (2.00, 4.00) | < 0.001 |
| Creatinine (μmol/L) | 0.8% | 72 (59, 86) | 84 (69, 117) | < 0.001 |
| Cystatin C (mg/L) | 3.6% | 1.38 (1.15, 1.81) | 1.81 (1.40, 2.47) | < 0.001 |
| DNR order | 0.4% | 3 (1.3%) | 26 (10%) | < 0.001 |
| H02AB | 5.4% | 15 (6.6%) | 30 (12%) | 0.035 |
| H03AA | 5.4% | 20 (8.8%) | 29 (12%) | 0.3 |
| IL-6 (ng/L) | 30.2% | 87 (40, 186) | 130 (62, 382) | < 0.001 |
| Leukocytes (x 10<sup>9</sup>/L) | 3.2% | 10 (7, 13) | 11 (8, 16) | 0.005 |
| M04AA | 5.4% | 14 (6.2%) | 25 (10%) | 0.11 |
| N02AA | 5.4% | 8 (3.5%) | 23 (9.5%) | 0.010 |
| N02BE | 5.4% | 41 (18%) | 56 (23%) | 0.2 |
| N03AX | 5.4% | 10 (4.4%) | 16 (6.6%) | 0.3 |
| N05BA | 5.4% | 10 (4.4%) | 15 (6.2%) | 0.4 |
| N05CF | 5.4% | 12 (5.3%) | 29 (12%) | 0.011 |
| N06AB | 5.4% | 12 (5.3%) | 17 (7.0%) | 0.4 |
| N06AX | 5.4% | 14 (6.2%) | 21 (8.6%) | 0.3 |
| Neutrophils (x 10<sup>9</sup>/L) | 21.3% | 8.2 (6.2, 10.9) | 9.2 (6.7, 13.8) | 0.002 |
| PPI medication | 5.4% | 42 (19%) | 62 (26%) | 0.067 |
| R03AC | 5.4% | 12 (5.3%) | 19 (7.8%) | 0.3 |
| R03AK | 5.4% | 16 (7.0%) | 15 (6.2%) | 0.7 |
| Y92BA | 5.4% | 17 (7.5%) | 26 (11%) | 0.2 |
| Y92AD | 5.4% | 13 (5.7%) | 23 (9.5%) | 0.13 |

Notes. Values are n (%) for categorical variables and median (Q1, Q3) for continuous variables.
<sup>1</sup> Group denominators shown in column headers. **Missing** indicates the percentage of missing observations per row.
<sup>2</sup> Pearson's $\chi^2$ , Fisher's exact, or Wilcoxon rank-sum test, as appropriate. Cells with $p < 0.05$ are grey-shaded.

The XGBoost survival model demonstrated acceptable internal validity, with a mean concordance index (C-index) of 0.74 (95% CI 0.73–0.75) on the full-case validation set and 0.76 (95% CI 0.74–0.77) on the training folds, indicating strong concordance between predicted and observed survival. The C-index remained stable until late in follow-up, after which a slight decrease was noted (Supplementary Table S2). Examination of Schoenfeld residuals confirmed the proportional hazards assumption for twelve of the thirteen most influential predictors of one-year mortality (Supplementary Figs. S1–S3). The assumption was violated for *number of medications per patient* (*p* = 0.01), consistent with a non-linear temporal effect of polypharmacy on survival risk.

### Feature importance and model explainability

Global model explainability using SHAP showed that age at admission was the dominant predictor of one-year mortality, followed by the number of medications per patient (Fig. 1). Polypharmacy ranked higher than both the Charlson Comorbidity Index (CCI) and the Clinical Frailty Scale (CFS), indicating that cumulative medication exposure captures vulnerability beyond conventional comorbidity or frailty metrics. Among ongoing pharmacotherapies, PPI use (ATC A02BC) was the only medication class included among the thirteen most influential predictors of mortality risk. ACEi displayed a small positive SHAP contribution, whereas ARB showed a slight negative effect (Fig. 2); however, the two antihypertensive classes ranked 72 and 59, respectively, of 87 variables and were therefore of low importance for risk prediction (Supplementary Table **??**).

**Figure 1.**
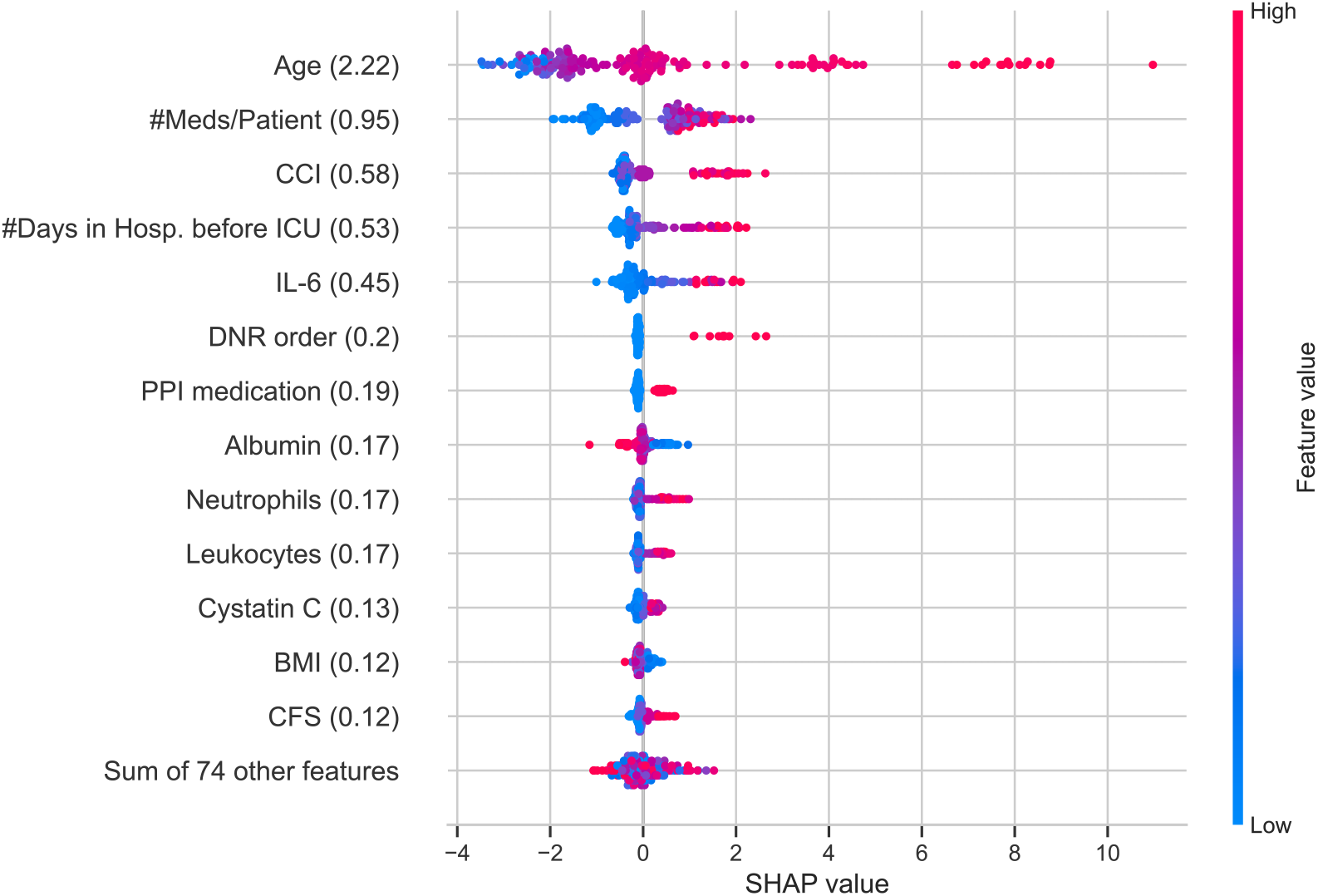
SHAP summary plot of the thirteen most influential variables in the XGBoost survival model (mean absolute SHAP values in parentheses). Higher SHAP values indicate a stronger contribution to predicted mortality risk. SHAP: SHapley Additive exPlanations; CCI: Charlson Comorbidity Index; ICU: Intensive Care Unit; IL-6: interleukin-6; DNR: Do Not Resuscitate; PPI: Proton Pump Inhibitors; BMI: Body Mass Index; CFS: Clinical Frailty Scale.

**Figure 2.**
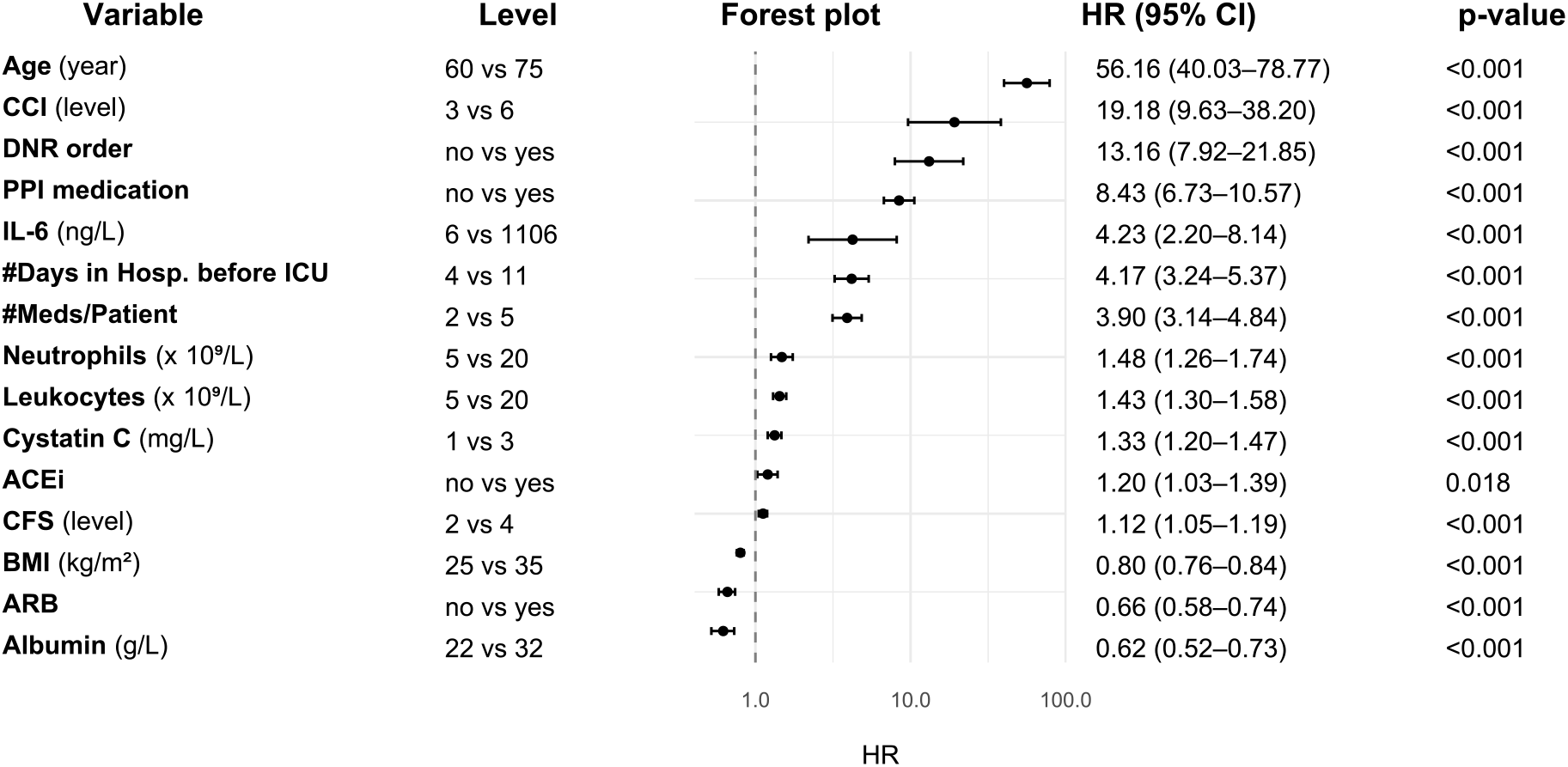
Bootstrapped SHAP-derived HRs with 95% CIs for the top thirteen predictors of one-year mortality, together with ACEi and ARB. Predictors are sorted by decreasing HR. Because the non-linear, tree-based model complicates traditional HR calculation, the additive SHAP framework enables surrogate derivations based on SHAP value importance. Accordingly, these HRs indicate the relative influence of a variable on predicted hazard for a given feature value rather than relative risks of mortality. For example, the hazard contribution of age is almost 60-fold higher for a 75-year-old compared with a 60-year-old. SHAP: SHapley Additive exPlanations; HR: Hazard Ratio; CI: Confidence Interval; ACEi: angiotensin-converting enzyme inhibitors; ARB: angiotensin II receptor blockers; CCI: Charlson Comorbidity Index; DNR: Do Not Resuscitate; PPI: Proton Pump Inhibitors; IL-6: interleukin-6; ICU: Intensive Care Unit; CFS: Clinical Frailty Scale; BMI: Body Mass Index.

Figure 1 summarises the global distribution of SHAP values for the thirteen top-ranking predictors. Variables with large positive SHAP values—such as advanced age, number of medications, CCI, days in hospital before ICU, and elevated IL-6—were associated with increased predicted mortality. These findings are consistent with general frailty, prolonged severe illness, and systemic inflammation as key drivers of mortality. In contrast, higher serum albumin was associated with negative SHAP values, likely reflecting better nutritional status and, by extension, lower frailty. BMI was also associated with negative SHAP values, although its importance was lower.

The forest plot (Fig. 2) displays bootstrapped SHAP-derived hazard ratios (HRs) with 95% confidence intervals (CIs) for the top thirteen predictors, together with ACEi and ARB. These HRs are not direct estimates of relative mortality risk; rather, they reflect the relative impact of each feature on the model’s predicted hazard across its observed range. Among the top predictors, advanced age was associated with by far the largest increases in predicted hazard, whereas higher serum albumin was associated with a reduced hazard contribution. The relative effect size for the number of medications per patient was comparable to, or larger than, that of major biomarkers of inflammation and renal function, emphasising the prognostic significance of pharmacological burden.

### Polypharmacy and survival

The dependency plot (Fig. 3) shows a clear monotonic increase in predicted mortality risk with rising medication count. The relationship was approximately linear up to four medications, after which the predicted hazard rose steeply, marking a threshold consistent with the conventional definition of polypharmacy (5 drugs). This accords with the non-proportionality observed in residual diagnostics and underscores a dose–response relationship between medication load and mortality risk.

**Figure 3.**
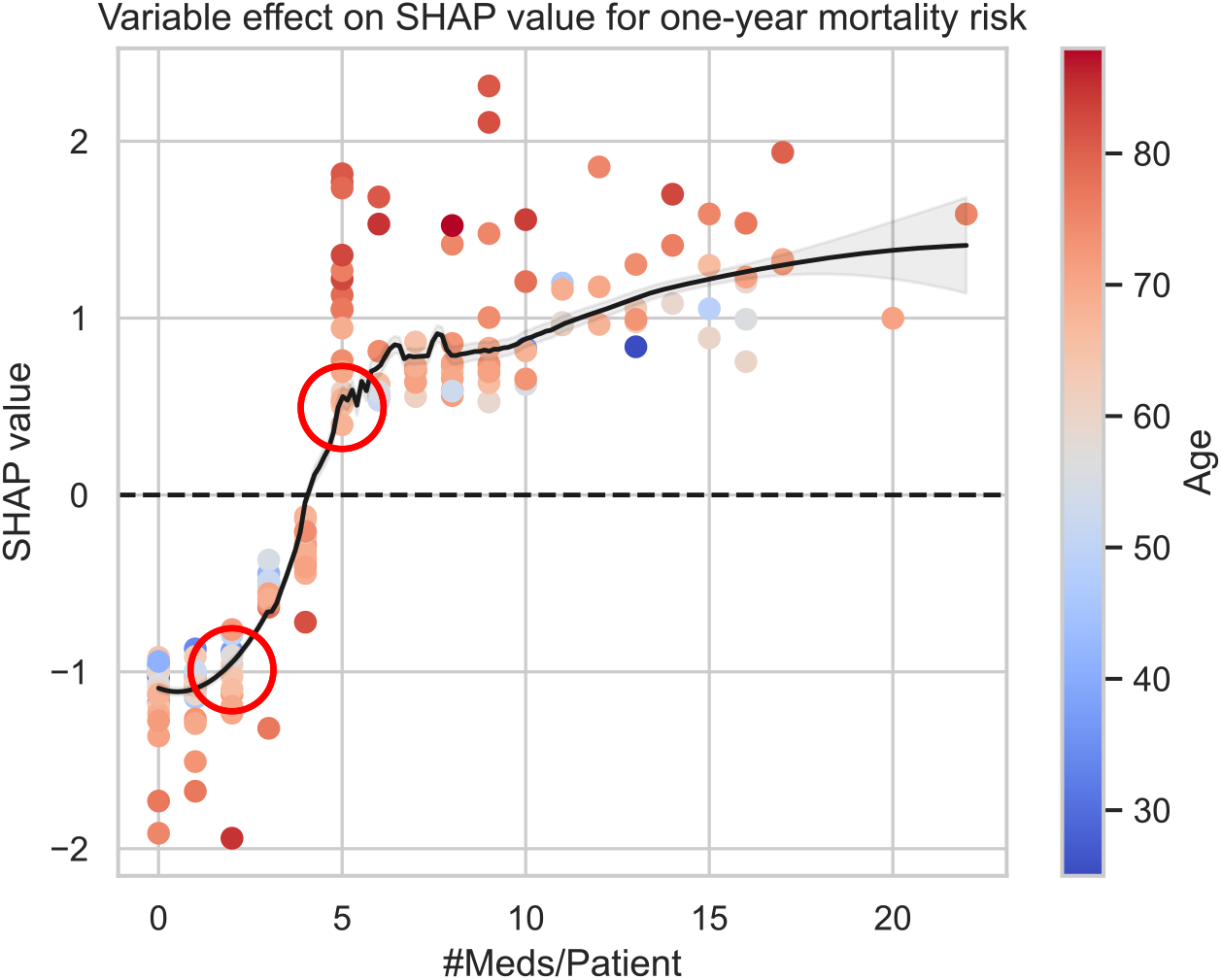
SHAP dependency plot illustrating the effect of the number of medications per patient on predicted mortality risk. Each point represents an individual patient and is coloured by age, from blue (younger) to red (older). Clinically selected reference values are highlighted with red circles. A steep hazard increase is observed at five or more concurrent medications, supporting the operational definition of polypharmacy. SHAP: SHapley Additive exPlanations.

Traditional survival analysis corroborated these findings. The Kaplan–Meier curves in Fig. 4 display the cumulative survival probability over one year stratified by polypharmacy status. Patients taking five or more medications had significantly lower survival than those taking four or fewer (log-rank *p <* 0.001). The separation of survival curves occurred early and persisted throughout follow-up, with some convergence towards the end of the year, suggesting late mortality among initially lower-risk patients.

**Figure 4.**
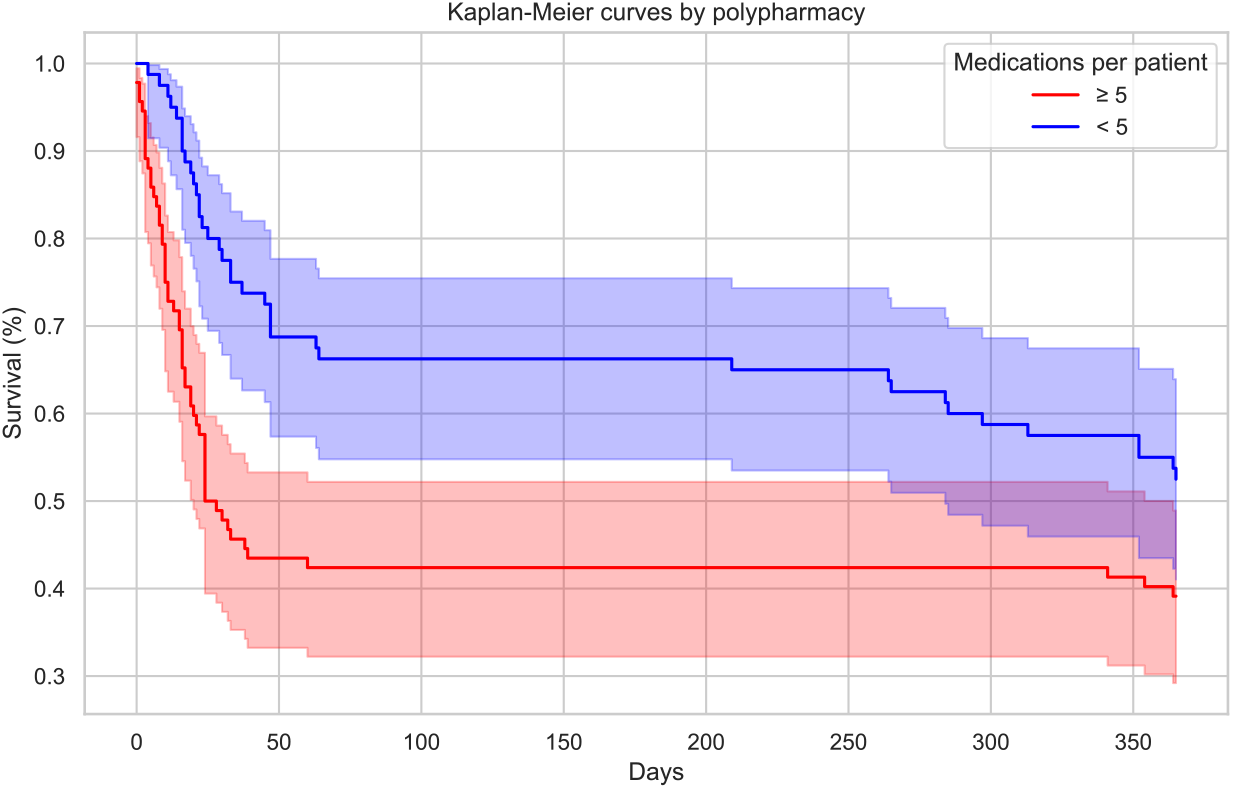
Kaplan–Meier survival curve stratified by polypharmacy status. Patients receiving five or more medications prior to ICU admission had significantly lower one-year survival than those taking four or fewer. ICU: Intensive Care Unit.

### Model robustness and clinical interpretation

Internal validation across repeated folds yielded stable feature rankings, indicating that the identified predictors were not driven by sampling variability. Model ensembling combined with risk estimation across multiple imputed datasets enhanced robustness to imputation-related uncertainty. Overall, the model consistently discriminated long-term outcomes in critical COVID-19.

## Discussion

Using a combined survival–machine learning framework in a large regional cohort with linked biobank data, we investigated the effect of ongoing outpatient medications on one-year mortality while jointly accounting for clinical physiology and inflammatory, endothelial, and renal biomarkers. We specifically evaluated ACEi and ARB given their putative mechanistic links to SARS-CoV-2 pathophysiology. In agreement with a recent meta-analysis in COVID-19^14^, our adjusted model found no evidence that ACEi or ARB exposure altered one-year mortality risk in the ICU setting. In contrast, we observed a strong, monotonic risk gradient with the *number of medications per patient* (Fig. 3): polypharmacy increased mortality independently of age, CCI, and CFS, suggesting that pharmacological load captures vulnerability beyond traditional comorbidity measures. This effect may generalise to broader ICU populations, but external validation is required.

Global explanation with SHAP supported and refined these observations. The SHAP summary (Fig. 1) showed that age, systemic inflammation (IL-6, neutrophils), and renal dysfunction (cystatin C) were dominant positive contributors to hazard, whereas higher serum albumin and BMI contributed negatively. This hierarchy reproduces the canonical pathobiology of fatal COVID-19 and mirrors the Swedish ICU experience described by Didriksson *et al*.^2^. Our model extends that framework by quantifying medication effects alongside these axes. Notably, our non-linear model assigned lower predictive importance to the number of admitted COVID-19 patients each day (a proxy for ICU burden) than previously reported, possibly because modelling non-linearity attenuates the apparent effect of ICU strain on mortality risk.

Among medications, PPIs (ATC A02BC) showed the largest positive SHAP contribution and an adjusted HR more than eight times higher for patients with ongoing therapy before ICU admission. Although biological rationales exist—hypochlorhydria, microbiome perturbation, and increased aspiration risk—the prior literature is inconsistent, with many associations attenuating after rigorous adjustment^9,10,15,16^. Given the strong correlation between PPI use, advanced age, and multimorbidity, residual confounding remains possible. However, our analyses incorporated comprehensive adjustment for age, comorbidity burden (CCI), frailty (CFS), and polypharmacy. To our knowledge, this breadth of adjustment has not been applied previously, and the persistence of the association suggests that PPI use may be independently associated with increased mortality in critical COVID-19. Smaller positive contributions for chronic glucocorticoids (H02AB) and thyroid replacement (H03AA) likely reflect baseline immune and metabolic vulnerability (Supplementary Table **??**). This does not contradict the established benefit of *in-hospital* dexamethasone for hypoxaemic COVID-19^17^. Conversely, modest negative SHAP values for DPP-4 inhibitors (A10BJ), inhaled LABA/ICS combinations (R03AK), dihydropyridine calcium-channel blockers (C08CA), and cardioselective *β*-blockers (C07AB) align with reports of neutral to slightly protective associations and plausible mechanisms (immune modulation, reduced airway viral-entry co-factors, endothelial stabilisation, and adrenergic control)^11–13,18–20^. Effect sizes were small and predictor rankings low; these findings should therefore be viewed as hypothesis-generating.

A methodological contribution of this work is a data-driven derivation of HRs for continuous and ordinal features that respects non-linearity. Rather than pre-specifying cut-points, we linked SHAP dependence functions to clinically chosen reference values, thereby using the full functional form learned by the model. Prior demonstrations of SHAP-derived effect contrasts used median splits and reported agreement with Cox coefficients^21^, but dichotomisation can mask clinically meaningful curvature. Our approach preserves shape information, improving interpretability at the bedside (e.g., diminishing returns of increasing PaO_2_ at higher ranges, or steep hazard inflexions for cystatin C above renal-injury thresholds).

Two findings merit emphasis for clinical translation. First, polypharmacy emerged as a high-impact, non-linear predictor: the proportional-hazards assumption was violated for this feature, and the Kaplan–Meier curves (Fig. 4) separated early between polypharmacy and non-polypharmacy groups. The dependency plot (Fig. 3) showed a sharp hazard escalation at ≥5 medications, reinforcing conventional operational definitions of polypharmacy^22^. Second, concordance with Didriksson *et al*.^2^ on the centrality of advanced age and low BMI strengthens confidence that our pharmacological insights are layered on a robust baseline risk structure rather than competing with it.

Strengths include a high-quality, multicentre registry and biobank linkage; careful curation; and an analysis plan integrating state-of-the-art survival learning with transparent explanations. Limitations are equally important. Simulated effect contrasts from a regularised, non-parametric model should be interpreted cautiously and validated externally; absolute risk impact depends on baseline risk (a modest HR can still be consequential at high baseline risk, and vice versa). IL-6 had substantial missingness, which we mitigated using multiple imputation; residual bias remains possible. Our exposure definition assumed that medications dispensed at least twice in the eight months before ICU admission represent ongoing therapy, which may misclassify intermittent or recently discontinued drugs. Finally, although we adjusted for comorbidity (CCI), frailty (CFS), and numerous biomarkers, confounding by indication and unmeasured severity cannot be fully excluded—particularly for PPI, chronic steroids, and mineral supplementation.

Overall, mortality after critical COVID-19 in this Swedish ICU cohort appears to be primarily determined by age, inflammation, renal function, and physiological derangement at admission, while pre-admission medication classes exert comparatively modest but interpretable effects. Polypharmacy, representing cumulative pharmacological exposure and frailty, emerged as a strong and independent predictor of poor survival—outperforming traditional comorbidity indices. PPI therapy contributed a small but significant positive hazard signal, though residual confounding is likely. These results complement previous analyses from the same SWECRIT cohort^2^, extending understanding of how chronic medication burden interfaces with acute pathophysiology and long-term mortality in critical COVID-19.

## Methods

### Study design and population

This retrospective study analysed prospectively and retrospectively collected data from critically ill adult patients (≥18 years) with laboratory-confirmed SARS-CoV-2 infection admitted to six ICUs in southern Sweden during the COVID-19 pandemic. The cohort and data infrastructure have been described in detail previously by Didriksson *et al*.^2^. Briefly, all participating ICUs contributed to the regional intensive care quality registry (*CovidIR*) and to the SWECRIT biobank, with structured recording of clinical variables, laboratory values, and treatment data. Exclusion criteria included ICU patients not admitted primarily for COVID-19 or those without obtained consent. For the present analysis, we included all patients with available one-year follow-up; individuals without confirmed follow-up were excluded. The study followed the WMA Declaration of Helsinki^23^ and was approved by the Swedish Ethical Review Authority (reference no. 2021-02323).

### Clinical data collection

Clinical and physiological data were obtained from the electronic medical record system, *CovidIR*, and the hospital patient administrative system for intensive care (PASIVA). Only variables available within ±1 hour of ICU admission were included to ensure temporal alignment with admission status. When admission data were missing, day 1 values were used as surrogates when available, including Sequential Organ Failure Assessment (SOFA) subscores. For a single case, an erroneous admission albumin value was replaced by the day 2 value verified in the source chart.

Where multiple observations existed within the inclusion window, laboratory variables were summarised by clinically meaningful extrema: bilirubin, D-dimer, C-reactive protein (CRP), ferritin, interleukin-6 (IL-6), creatinine, lactate, leukocytes, neutrophils, and procalcitonin (PCT) were recorded at their *maximum*, whereas lymphocytes, arterial oxygen tension (PaO_2_), PF quotient, arterial blood pH, white blood cell count, and arterial carbon dioxide tension (PaCO_2_) were taken at their *minimum*. The PF quotient was defined as the ratio of PaO_2_ to inspired oxygen fraction (FiO_2_). The Glasgow Coma Scale (GCS) was derived from the Reaction Level Scale 1985 (RLS85) using standard conversion. Invasive mechanical ventilation (IMV) was coded as present if initiated within the first 24 hours of ICU admission, accommodating minor documentation delays.

Prescription data were retrieved from the Swedish National Prescribed Drug Register, capturing all pharmacy dispensations nationwide. To ensure inclusion of ongoing therapies rather than sporadic prescriptions, drugs were considered *active* if dispensed at least twice in the eight months preceding ICU admission. Medications were classified at level 4 of the Anatomical Therapeutic Chemical (ATC) system, and the 25 most common prescriptions among survivors, non-survivors, and the total population were analysed alongside the overall number of medications per patient. ACEi and ARB were analysed as separate exposures. Polypharmacy was defined as the use of *≥* 5 concurrent medications^22^.

### Outcome definition

The primary outcome was time to death from ICU admission, with administrative censoring at 365 days. Surviving patients were censored at one year. The time origin (day 0) corresponded to ICU admission, ensuring comparability with the one-year outcome framework used in Didriksson *et al*.^2^.

### Handling of missing data

Variables with *>* 20% missing data were excluded, except for IL-6 (30.2%), mean arterial pressure (20.3%), and the PF quotient (37.6%), which were retained due to established clinical relevance in critical illness. Binary variables with *<* 2% prevalence in one category were removed to avoid model imbalance. Remaining missing values were imputed using Multiple Imputation by Chained Equations (MICE)^24^ via the `miceforest` algorithm (version 5.6.2), which iteratively performs random-forest imputations until convergence. We created ten imputed datasets. True and imputed variable-distribution plots were inspected to confirm plausibility.

### Model development and validation

We implemented an XGBoost survival model using a Cox partial-likelihood objective (`XGBoost`, version 2.1.2), which generalises the proportional hazards model to handle non-linear and interaction effects via gradient-boosted trees^25^. One model was trained per imputed dataset, and final risk predictions were calculated as the mean across models. Hyperparameter tuning used the `Optuna` tree-structured Parzen estimator (version 3.6.0)^26^. A repeated stratified 5-fold cross-validation scheme (20 repeats, 100 tuning rounds) optimised performance based on mean internal-validation C-index. Optimal hyperparameters were: maximum tree depth 2, learning rate 0.00411, subsample fraction 0.652, minimum split loss 0.339, minimum child weight 1, L1 (alpha) 0.345, and L2 (lambda) 1.297. Computations were performed on a Windows workstation (Intel Core i7-8700K, 3.70 GHz) using Python 3.12.7.

Model calibration was inspected using time-dependent C-indices across 90-, 180-, 270-, and 365-day horizons to verify that learned hazards remained proportionate over time. Schoenfeld residuals of SHAP values were used to evaluate the proportional hazards assumption. Full-case patients (those without imputation) were reserved for internal validation to minimise potential imputation bias.

### Model interpretability

Model interpretability was assessed using SHAP (version 0.46.0)^27^. SHAP values were computed from risk predictions on the internal validation set, comprising data not used during training. Global importance was visualised via a summary plot (Fig. 1), ranking predictors by absolute mean SHAP value. Non-linear effects were examined using SHAP dependency plots, visualising marginal associations between individual feature values and predicted log-hazard. To approximate HRs from SHAP outputs, we used bootstrap resampling of non-overlapping subsamples to estimate mean differences in SHAP values between contrasting exposure states, scaled by feature-specific standard deviations. For binary variables, the HR corresponds to the contrast between presence and absence of exposure. For continuous and ordinal variables, clinically meaningful reference points were defined based on domain expertise and the shape of dependency plots, ensuring interpretability within physiological ranges. The resulting HRs with 95% CIs were visualised in a forest plot (Fig. 2).

### Statistical considerations

All tests were two-sided with a significance level of *p <* 0.05. No *a priori* power calculation was performed, as the study was exploratory and included all eligible ICU admissions during the study period. Non-response and missingness characteristics have been reported previously^2^. Statistical analyses were performed using Python 3.12.7 and R 4.4.2.

## Code availability

The code is available at https://github.com/malinhjartstrom/polypharmacy.

## Data availability

The data supporting the findings of this study are not publicly available due to ethical and legal restrictions, but can be obtained from A.F. upon reasonable request.

## Acknowledgements

We thank the staff at Skåne University Hospital in Malmö and Lund, Helsingborg Hospital, and Kristianstad Hospital for their valuable support in data collection.

## Author contributions statement

A.F. and M.H. designed the study. I.D. and M.S. collected clinical data. M.H. performed the statistical analyses and prepared figures and tables, in dialogue with A.F. and A.J. A.F. and M.H. wrote the initial manuscript. All authors reviewed the manuscript and approved the final version.

## Additional information

The authors declare no competing interests.

