## Supplemental tables and figures for "Polypharmacy and Proton Pump Inhibitor Use Independently Predict One-Year Mortality in Critical COVID-19: An Explainable AI–Based Survival Analysis"

### SUPPLEMENTAL INFORMATION

**Table S1.** Patient characteristics stratified by one-year mortality from ICU admission

| Characteristic | Missing | One-year mortality | | p-value[2] |
|---|---|---|---|---|
| | | **Survivors** (N = 235)[1] | **Non-survivors** (N = 262)[1] | |
| **# Days in hospital before ICU** | 2.0% | 2.0 (0.0, 5.0) | 3.0 (1.0, 7.0) | < 0.001 |
| **# Meds/Patient** | 5.4% | 3.0 (1.0, 7.0) | 5.0 (2.0, 10.0) | < 0.001 |
| **# Survival days from ICU admission** | 0.0% | 365 (365, 365) | 19 (10, 113) | < 0.001 |
| **# Symptomatic days** | 1.0% | 8.0 (6.0, 10.0) | 7.0 (5.0, 10.0) | 0.021 |
| **A10BA** | 5.4% | 42 (19%) | 38 (16%) | 0.4 |
| **A10BK** | 5.4% | 9 (4.0%) | 10 (4.1%) | >0.9 |
| **A10BJ** | 5.4% | 11 (4.8%) | 14 (5.8%) | 0.7 |
| **A12AX** | 5.4% | 8 (3.5%) | 25 (10%) | 0.004 |
| **ACEi** | 5.4% | 48 (21%) | 43 (18%) | 0.3 |
| **Age** (years) | 0.0% | 62 (53, 68) | 70 (61, 75) | < 0.001 |
| **Albumin** (g/L) | 3.6% | 27.8 (25.2, 29.9) | 27.3 (24.8, 30.0) | 0.2 |
| **ARB** | 5.4% | 31 (14%) | 61 (25%) | 0.002 |
| **Arterial oxygen tension** (kPa) | 16.9% | 8.80 (7.50, 10.10) | 8.80 (7.70, 10.20) | 0.8 |
| **Arterial pH** | 3.0% | 7.44 (7.37, 7.47) | 7.42 (7.35, 7.46) | 0.027 |
| **B01AC** | 5.4% | 31 (14%) | 66 (27%) | < 0.001 |
| **B01AF** | 5.4% | 10 (4.4%) | 19 (7.8%) | 0.12 |
| **B03BA** | 5.4% | 23 (10%) | 27 (11%) | 0.7 |
| **B03BB** | 5.4% | 4 (1.8%) | 18 (7.4%) | 0.004 |
| **Bilirubin** (μmol/L) | 4.6% | 8.0 (6.0, 11.0) | 9.0 (6.0, 13.0) | 0.007 |
| **BMI** ($kg/m^2$) | 2.4% | 31 (27, 35) | 29 (26, 34) | 0.016 |
| **Body temperature** (°C) | 3.2% | 37.50 (37.00, 38.10) | 37.40 (36.80, 38.00) | 0.056 |
| **C01DA** | 5.4% | 7 (3.1%) | 14 (5.8%) | 0.2 |
| **C03CA** | 5.4% | 14 (6.2%) | 32 (13%) | 0.011 |
| **C07AB** | 5.4% | 52 (23%) | 77 (32%) | 0.033 |

*Continued on next page*

**Table S1** (continued): Patient characteristics by one-year mortality

| Characteristic | Missing | One-year mortality | | p-value[2] |
|---|---|---|---|---|
| | | **Survivors** (N = 235)[1] | **Non-survivors** (N = 262)[1] | |
| **C08CA** | 5.4% | 54 (24%) | 59 (24%) | > 0.9 |
| **C10AA** | 5.4% | 51 (22%) | 85 (35%) | 0.003 |
| **Calprotectin** ($\mu$g/g) | 3.6% | 7 (4, 11) | 8 (5, 13) | 0.009 |
| **Cancer** | 0.0% | 25 (11%) | 49 (19%) | 0.012 |
| **Cardiovascular MAP** (mmHg) | 20.3% | 70 (65, 80) | 71 (65, 80) | > 0.9 |
| **CCI** | 0.0% | 2.00 (1.00, 3.00) | 3.00 (2.00, 4.00) | < 0.001 |
| **Cerebrovascular insult** | 0.0% | 7 (3.0%) | 27 (10%) | 0.001 |
| **CFS** | 0.8% | 3.00 (2.00, 3.00) | 3.00 (2.00, 4.00) | < 0.001 |
| **Chronic kidney disease** | 0.0% | 5 (2.1%) | 15 (5.7%) | 0.042 |
| **Chronic pulmonary disease** | 0.0% | 45 (19%) | 49 (19%) | 0.9 |
| **Congestive heart failure** | 0.0% | 14 (6.0%) | 30 (11%) | 0.031 |
| **Creatinine** ($\mu$mol/L) | 0.8% | 72 (59, 86) | 84 (69, 117) | < 0.001 |
| **CRP** | 0.8% | 155 (89, 222) | 151 (83, 234) | 0.7 |
| **Cystatin C** (mg/L) | 3.6% | 1.38 (1.15, 1.81) | 1.81 (1.40, 2.47) | < 0.001 |
| **D-dimer** ($\mu$g/mL) | 12.1% | 2 (1, 4) | 2 (1, 8) | 0.012 |
| **Diabetes mellitus (complicated)** | 0.0% | 27 (11%) | 44 (17%) | 0.092 |
| **Diabetes mellitus (uncomplicated)** | 0.0% | 41 (17%) | 41 (16%) | 0.6 |
| **DNR order** | 0.4% | 3 (1.3%) | 26 (10%) | < 0.001 |
| **Endostatin** (ng/L) | 3.6% | 58,662 (46,891, 73,508) | 66,264 (52,702, 90,185) | < 0.001 |
| **Ferritin** (ng/mL) | 15.5% | 1,427 (669, 2,229) | 1,432 (929, 2,440) | 0.2 |
| **Glasgow Coma Scale** | 1.8% | 5.00 (5.00, 14.00) | 5.00 (5.00, 14.00) | 0.2 |
| **H02AB** | 5.4% | 15 (6.6%) | 30 (12%) | 0.035 |
| **H03AA** | 5.4% | 20 (8.8%) | 29 (12%) | 0.3 |
| **Habitual creatinine** ($\mu$mol/L) | 9.5% | 76 (65, 90) | 81 (73, 98) | < 0.001 |
| **Heart rate** (beats/min) | 2.8% | 90 (80, 106) | 92 (80, 110) | 0.2 |
| **Hypertension** | 0.8% | 119 (51%) | 152 (58%) | 0.10 |
| **ICAM** | 3.8% | 279,451 (217,486, 349,602) | 299,433 (232,849, 372,126) | 0.028 |
| **ICU burden** | 0.0% | 27 (18, 39) | 34 (23, 55) | < 0.001 |
| **IL-6** (ng/L) | 30.2% | 87 (40, 186) | 130 (62, 382) | < 0.001 |
| **IMV within 24 h of ICU admission** | 1.0% | 115 (50%) | 158 (61%) | 0.017 |
| **Lactate** (mmol/L) | 2.6% | 2.10 (1.60, 2.50) | 2.40 (1.80, 3.45) | < 0.001 |
| **Leukocytes** ($\times10^9$/L) | 3.2% | 10 (7, 13) | 11 (8, 16) | 0.005 |
| **Lymphocytes** ($\times10^3/\mu$L) | 21.7% | 0.60 (0.45, 0.90) | 0.60 (0.40, 0.90) | 0.13 |
| **M04AA** | 5.4% | 14 (6.2%) | 25 (10%) | 0.11 |
| **Myocardial infarction** | 0.0% | 22 (9.4%) | 36 (14%) | 0.13 |
| **N02AA** | 5.4% | 8 (3.5%) | 23 (9.5%) | 0.010 |
| **N02BE** | 5.4% | 41 (18%) | 56 (23%) | 0.2 |
| **N03AX** | 5.4% | 10 (4.4%) | 16 (6.6%) | 0.3 |
| **N05BA** | 5.4% | 10 (4.4%) | 15 (6.2%) | 0.4 |
| **N05CF** | 5.4% | 12 (5.3%) | 29 (12%) | 0.011 |
| **N06AB** | 5.4% | 12 (5.3%) | 17 (7.0%) | 0.4 |
| **N06AX** | 5.4% | 14 (6.2%) | 21 (8.6%) | 0.3 |
| **Neutrophils** ($\times10^9$/L) | 21.3% | 8.2 (6.2, 10.9) | 9.2 (6.7, 13.8) | 0.002 |
| **NGAL** | 3.6% | 98,783 (75,211, 141,693) | 133,291 (95,212, 197,119) | < 0.001 |
| **NOAK** | 0.0% | 5 (2.1%) | 11 (4.2%) | 0.2 |
| **Noradrenaline** ($\mu$g/kg/min) | 12.9% | | | 0.029 |
| 0 | | 151 (74%) | 141 (62%) | |
| $\leq 0.1$ | | 34 (17%) | 51 (22%) | |
| $> 0.1$ | | 20 (9.8%) | 36 (16%) | |
| **$PaCO_2$** (kPa) | 5.8% | 4.40 (4.00, 4.90) | 4.40 (3.90, 5.10) | > 0.9 |
| **$PaO_2$** (kPa) | 6.0% | 7.30 (6.50, 8.30) | 7.40 (6.60, 8.10) | > 0.9 |
| **$PaO_2/FiO_2$ ratio** (kPa) | 6.6% | 12 (9, 18) | 11 (8, 17) | 0.14 |

*Continued on next page*

**Table S1** (continued): Patient characteristics by one-year mortality

| Characteristic | Missing | One-year mortality | | p-value[2] |
|---|---|---|---|---|
| | | **Survivors** (N = 235)[1] | **Non-survivors** (N = 262)[1] | |
| **Peptic ulcer disease** | 0.0% | 2 (0.9%) | 8 (3.1%) | 0.11 |
| **Peripheral vascular disease** | 0.0% | 10 (4.3%) | 14 (5.3%) | 0.6 |
| **PPI medication** | 5.4% | 42 (19%) | 62 (26%) | 0.067 |
| **Procalcitonin** (ng/mL) | 15.3% | 0.4 (0.2, 1.0) | 0.5 (0.3, 1.4) | 0.002 |
| **R03AC** | 5.4% | 12 (5.3%) | 19 (7.8%) | 0.3 |
| **R03AK** | 5.4% | 16 (7.0%) | 15 (6.2%) | 0.7 |
| **Rheumatic disease** | 0.0% | 19 (8.1%) | 22 (8.4%) | 0.9 |
| **Sex** | 0.0% | | | 0.7 |
| Female | | 64 (27%) | 67 (26%) | |
| Male | | 171 (73%) | 195 (74%) | |
| **Smoker** | 0.6% | | | 0.003 |
| No, never | | 148 (63%) | 126 (48%) | |
| Yes, previously | | 77 (33%) | 116 (45%) | |
| Yes, presently | | 9 (3.8%) | 18 (6.9%) | |
| **Systolic blood pressure** (mmHg) | 2.2% | 120 (107, 138) | 115 (99, 130) | 0.002 |
| **VCAM** | 3.6% | 464,527 (358,082, 647,564) | 551,085 (418,057, 728,324) | $< 0.001$ |
| **WBC** ($\times 10^9$/L) | 3.4% | 269 (191, 361) | 249 (180, 329) | 0.019 |
| **Y92BA** | 5.4% | 17 (7.5%) | 26 (11%) | 0.2 |
| **Y92AD** | 5.4% | 13 (5.7%) | 23 (9.5%) | 0.13 |

*Notes.* Values are *n* (%) for categorical variables and median (Q1, Q3) for continuous variables.
[1] Group denominators are shown in column headers. **Missing** indicates the percentage of missing observations per row.
[2] Pearson's $\chi^2$, Fisher's exact, or Wilcoxon rank-sum test, as appropriate. Cells with $p < 0.05$ are grey-shaded.

**Table S2.** C-index at 90, 180, 270, and 365 days from ICU admission

| Time (days) | C-index (95% CI) |
|---|---|
| 90 | 0.77 (0.76–0.79) |
| 180 | 0.77 (0.75–0.80) |
| 270 | 0.77 (0.75–0.78) |
| 365 | 0.74 (0.73–0.75) |

*Notes.* C-indices with 95% confidence intervals (CIs) were computed using risk predictions from internal validation. Overall performance remained stable, with a minor decline during the last quarter of the year. C-index: concordance index; CI: confidence interval.

**Table S3.** Adjusted SHAP-based hazard ratios for one-year mortality

| Ongoing medication | HR (95% CI) | p-value | Survival model ranking |
|---|---|---|---|
| A02BC | 8.43 (6.73–10.57) | <0.001 | 7 |
| A12AX | 4.03 (2.59–6.27) | <0.001 | 28 |
| C08CA | 0.30 (0.23–0.40) | <0.001 | 37 |
| N03AX | 0.34 (0.26–0.44) | <0.001 | 39 |
| C07AB | 0.46 (0.36–0.58) | <0.001 | 49 |
| R03AK | 0.49 (0.40–0.61) | <0.001 | 51 |
| H03AA | 1.60 (1.27–2.02) | <0.001 | 52 |
| H02AB | 1.79 (1.43–2.24) | <0.001 | 53 |
| A10BJ | 0.50 (0.43–0.59) | <0.001 | 54 |
| Y92BA | 0.49 (0.41–0.57) | <0.001 | 55 |
| C10AA | 1.29 (1.04–1.61) | 0.018 | 57 |

*Continued on next page*

**Table S3** (continued): Adjusted SHAP-based HRs for one-year mortality

| **Ongoing medication** | **HR (95% CI)** | **p-value** | **Survival model ranking** |
|---|---|---|---|
| B03BA | 1.53 (1.28–1.83) | <0.001 | 58 |
| B01AC | 1.16 (0.91–1.48) | 0.236 | 59 |
| ARB | 0.66 (0.58–0.74) | <0.001 | 60 |
| C03CA | 1.43 (1.05–1.95) | 0.020 | 63 |
| R03AC | 1.31 (0.91–1.88) | 0.144 | 65 |
| N06AX | 0.66 (0.54–0.82) | <0.001 | 66 |
| N05CF | 1.11 (0.76–1.61) | 0.589 | 67 |
| M04AA | 0.73 (0.57–0.92) | 0.008 | 69 |
| A10BA | 0.77 (0.66–0.90) | 0.001 | 71 |
| ACEi | 1.20 (1.03–1.39) | 0.018 | 73 |
| Y92AD | 0.80 (0.65–0.98) | 0.031 | 74 |
| A10BK | 0.73 (0.55–0.95) | 0.019 | 75 |
| N02AA | 1.17 (0.96–1.43) | 0.105 | 78 |
| B03BB | 0.89 (0.70–1.12) | 0.322 | 80 |
| N02BE | 1.11 (0.96–1.30) | 0.158 | 82 |
| C01DA | 0.91 (0.77–1.07) | 0.245 | 84 |
| N05BA | 1.06 (0.92–1.23) | 0.404 | 85 |
| B01AF | 0.94 (0.82–1.08) | 0.377 | 86 |
| N06AB | 0.96 (0.87–1.05) | 0.363 | 87 |

*Notes.* Adjusted hazard ratios (HRs) with 95% CIs were derived from SHAP contrasts in the XGBoost survival model. Ongoing medications are listed in order of decreasing importance for risk prediction. A02BC is the level 4 ATC code for PPI medication. Variables significant at $p < 0.05$ are grey-shaded. HR: hazard ratio; CI: confidence interval; PPI: proton pump inhibitor.

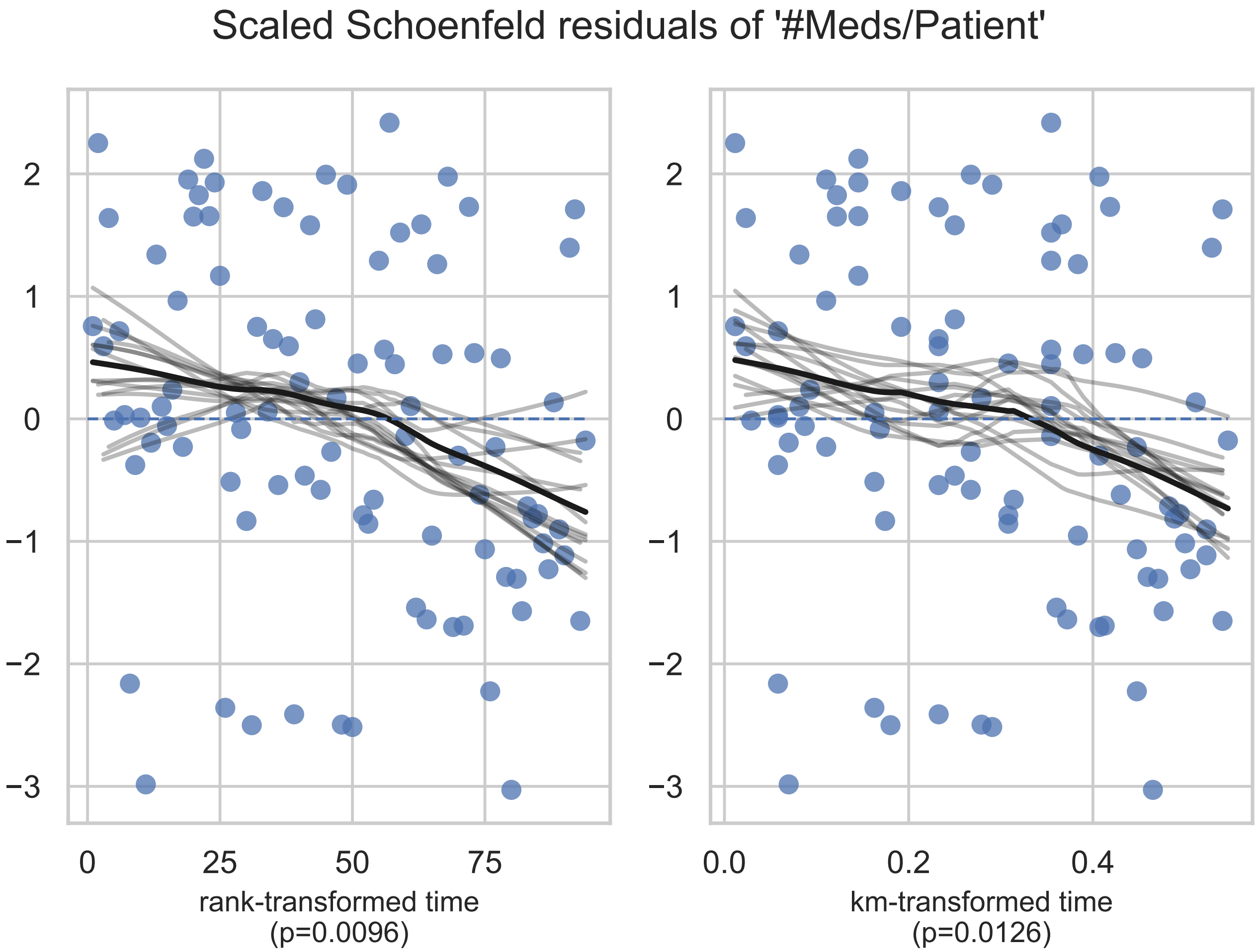


**Figure S1.** Schoenfeld residuals for *number of medications per patient* showing violation of the proportional-hazards assumption ($p < 0.01$).

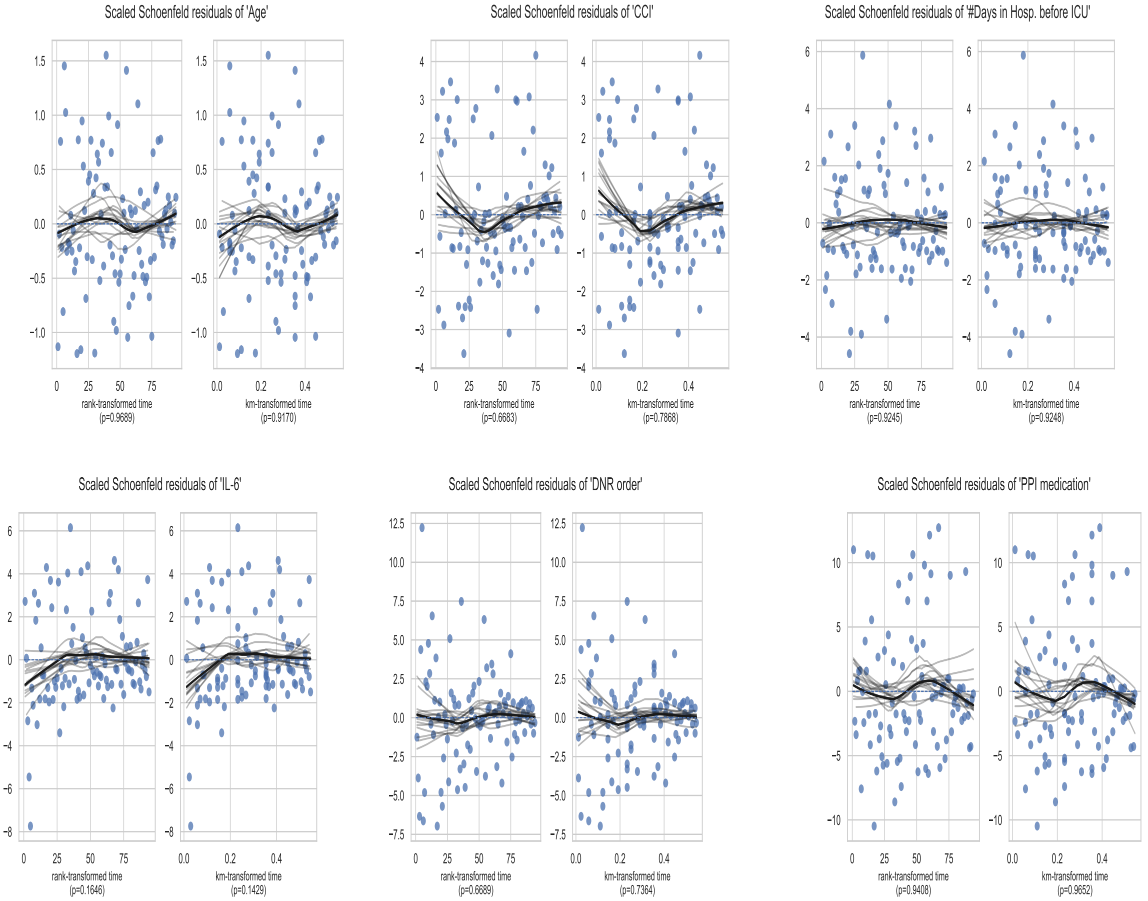


**Figure S2.** Schoenfeld residuals for age, CCI, number of days in hospital before ICU, IL-6, DNR order, and PPI medication. CCI: Charlson Comorbidity Index; IL-6: interleukin-6; DNR: Do Not Resuscitate; PPI: proton pump inhibitors.

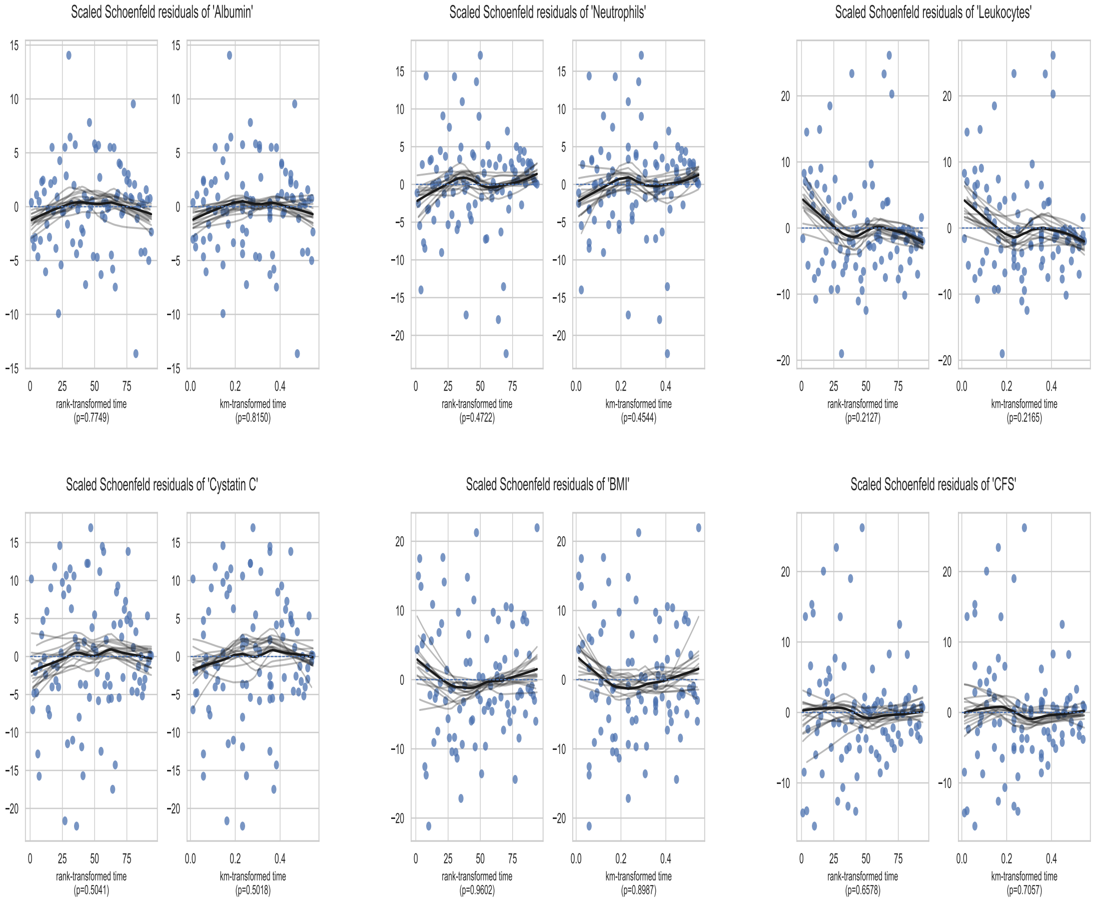


**Figure S3.** Schoenfeld residuals for albumin, neutrophils, leukocytes, cystatin C, BMI, and CFS. BMI: body mass index; CFS: Clinical Frailty Scale.
